# Microvascular Metrics on Diabetic Retinopathy: Insights from a Meta-Analysis of Diabetic Eye Images from Real-World Data

**DOI:** 10.1101/2024.08.01.24311332

**Authors:** Cristina Cuscó, Pau Esteve, Ana Almazán-Moga, Jimena Fernández-Carneado, Berta Ponsati

## Abstract

**Objective:** To quantify microvascular lesions in a large Real-World Data (RWD) set, based on single central retinal fundus images from different origins, with the aim of validating its use as a precision tool for classifying Diabetic Retinopathy (DR) severity.

**Design:** Retrospective meta-analysis across multiple fundus image datasets.

**Sample size:** The study analyzed 2,340 retinal fundus images from diabetic patients across four diverse RWD international datasets, including populations from Spain, India, China and the US.

**Intervention:** The quantification of specific microvascular lesions: microaneurysms (MAs), hemorrhages (Hmas) and hard exudates (HEs) using advanced automated image analysis techniques on central retinal images to validate reliable metrics for DR severity assessment. The images were pre-classified in the DR severity levels as defined by the International Clinical Diabetic Retinopathy (ICDR) scale.

**Main Outcome Measures:** The primary variables measured were the number of MAs, Hmas, red lesions (RLs) and HEs. These counts were related with DR severity levels using statistical methods to validate the relationship between lesion counts and disease severity.

**Results:** The analysis revealed a robust and statistically significant increase (p<0.001) in the number of microvascular lesions and the DR severity across all datasets. Tight data distributions were reported for MAs, Hmas and RLs, supporting the reliability of lesion quantification for accurately assessing DR severity. HEs also followed a similar pattern, but with a broader dispersion of data. Data used in this study are consistent with the definition of the DR severity levels established by the ICDR guidelines.

**Conclusions:** The statistically significant increase in the number of microvascular lesions across DR severity validate the use of lesion quantification in a single central retinal field as a key biomarker for disease classification and assessment. This quantification method demonstrates an improvement over traditional assessment scales, providing a quantitative metric that enhances the precision of disease classification and patient monitoring. The inclusion of a numerical component allows for the detection of subtle variations within the same severity level, offering a deeper understanding of disease progression. The consistency of results across diverse datasets not only confirms the method’s reliability but also its applicability in a global healthcare setting.

## 1. Introduction

Diabetic Retinopathy (DR) emerges as a silent yet significant threat to vision, caused by diabetes mellitus as the underlying cause. ^1–3^ It orchestrates a complex sequence of events in the retinal microvascular network, where chronic hyperglycemia triggers a cascade of microvascular lesions that begin with microaneurysms (MAs) and continue with hemorrhages (Hmas) and exudates (HEs).^4,5^ These microvascular abnormalities not only determine the onset of DR, but their close examination has proved to reveal the disease severity and risk progression. ^6–8^ Particularly, the turnover rate and number of MAs have been identified as significant predictors of disease progression, meaning that high counts correlate with rapid advancement to more severe stages and/or precipitation to central-involved macular edema.^9–13^

Globally, the assessment and management of DR rely on fundus photography, a technique that revolutionized the way DR severity is classified, by documenting and monitoring retinal changes over time.^14–16^ This method provides quantifiable means of assessing retinal alterations across various disease stages.^17^ The precise capture and annotation of several (3 to 7) retinal images enable detailed analysis of microvascular lesions, leading to an accurate and standardized classification.^18^ In our recent study, we explored the potential of quantifying microvascular lesions within a single central retinal fundus photography as a predictive marker for DR severity. By precisely counting microaneurysms (MAs), hemorrhages (Hmas), and hard exudates (HEs) in a single image across different NPDR stages, we established a robust relationship, with high statistical significance, between the number of lesions and the severity of DR. ^19^

These findings offer a substantial value not only in DR diagnostics, but specially in clinical trials, since it enables a more nuanced and quantitative differentiation, even within the same DR severity level. This method marks an improvement over DR severity sub-classification systems such as ETDRS scale, which, despite being the regulatory standard for DR evaluation in clinical trials, lacks quantitative basis, as the assessments rely on qualitative observations.^20^ Furthermore, the ETDRS system is likely to become outdated as fundus imaging techniques evolve, offering more detailed views of the retina within a single image. ^21–23^ The International Classification of Diabetic Retinopathy (ICDR) system also has its own limitations: its classification categories (no DR, mild, moderate and severe NPDR) are quite broad, so the spectrum of conditions falling in-between these levels are not distinctly recognized and are treated as equivalent despite noticeable differences.

Our approach highlights the potential of using robust numerical biomarkers to classify the DR stage, an option not presently available with existing methodologies. Interestingly, it also simplifies the assessment process required by the ETDRS protocol from seven fields to only one, and determines with high accuracy the severity level of the patient through their microvascular state. On the overall, these advantages directly impact on the personalization of treatment strategies and the enhancement of precision medicine.

Owing to the accessibility of public fundus photography databases, a comprehensive characterization of the microvascular component of diabetic retinopathy (DR) has become achievable across a diverse demographic spectrum. In this regard, an extensive collection of images varying in nature, origin and quality, previously classified according to the International Classification Diabetic Retinopathy (ICDR) on mild, moderate and severe DR, has served as a rich source for in-depth quantifiable analysis and the basis for a more accurate and standardized patient categorization. Building on this framework, our current study aims to validate, in a broad and representative sample of Real-World Data, the predictive utility of quantifying retinal lesions within a single retinal central field as a precise metric for determining the severity of DR. The confirmation of this approach in a broad sample of data seeks to enlarge the evidence of having numerical biomarkers as a powerful tool for precise DR severity assessment, resulting in improved clinical outcomes and patient care.

## 2. Methods

### 2.1 Datasets

In this retrospective image analysis study, four independent retinal image datasets were included to assess the relationship between ICDR grade and microvascular lesion number. All datasets contained original fundus images and annotated images for segmentation purposes.

Dataset 1 (n=144) is a private dataset representative of Spanish population. It has already been discussed in a recent publication to preliminarily test the stated hypothesis.^19^ The acquisition protocol was performed in the NM-I field with a 45° fundus camera under mydriasis with tropicamide 0.5%.

Dataset 2 (n=586) is also publicly available and representative of Chinese population. Fundus images were captured using a 45° fundus centered at middle distance between the optic disc and the fovea.^24^

Dataset 3 (n=1,395) is available on-request and consists of lesion-segmented images derived from the original EyePACS dataset,^25^ mainly representative of the Chinese and US population. Fundus images are centered in the fovea or nearby using a 45° camera.

Finally, Dataset 4 (n=50) is also publicly available, representative of Indian population. The images were acquired centered on the fovea using a 50° fundus camera. All the subjects in the dataset had undergone prior mydriasis.^26^

The publication of the image sets was authorized by the corresponding organizations before making them available. Informed consent was not required for this retrospective study, except for Dataset 1, which required a signed informed consent by the participants. The study adhered to the tenants of the Declaration of Helsinki.

### 2.2 Image segmentation and counting

The image lesion annotations and DR gradings from all datasets was manually performed or supervised by retina specialists prior to this work. ^19,24–26^

In the present study, the retinal lesions were automatically counted based on the segmented image files provided, using a connected components analysis method on the binary images. The implementation of a watershed algorithm aided in discerning overlapping lesions. Robustness of this counting method was evaluated and tuned using images containing XML files with individual lesion mappings for comparison.

Resulting data was filtered out to discard images with heavy outlier counts (1.5×IQR more unusual than Q1 or Q3) for MAs and Hmas from the datasets:

Dataset 1: total data: n=160 / filtered data: n=144

Dataset 2: total data: n=681/ filtered data: n=586

Dataset 3: total data: n=1,549/ filtered data: n=1,395

Dataset 4: total data: n=55/ filtered data: n=50.

Intergroup differences between DR severity levels were evaluated individually within each set using two-sided unpaired Wilcoxon signed-rank tests (α=0.05).

For the supplementary analysis of a subset having a severe classification with a low MA count (n=37), the Check Eye platform was used. Check Eye is a cloud-based solution to detect DR and specific lesions using fundus images through a convolutional neural network. (exact citation pending).

### 2.3 Prediction model

To assess the DR level predictability from the multiple lesion counts, a classification neural network with layer neuron size 7 and weight decay 0.001 was set up using caret R package. Values were scaled and merged, 80 % of the total samples were used for training and the remaining 20% for testing.

## 3. Results

Firstly, the number of MAs, Hmas and red lesions (RLs), understood as a combination of MAs and Hmas, was quantified across four severity stages (no DR, mild NPDR, moderate NPDR and severe NPDR) in each of the four databases. Additionally, the count for HEs was also performed.

A total of 2,175 retinal fundus images from diabetic patients were included in the analysis, although the rough sample (n=2,340) was also used for comparison purposes. The number of MAs, Hmas and RLs quantified from a single segmented central retinal field were plotted against ICDR severity levels, as shown in Figure 1:

**Fig. 1.**
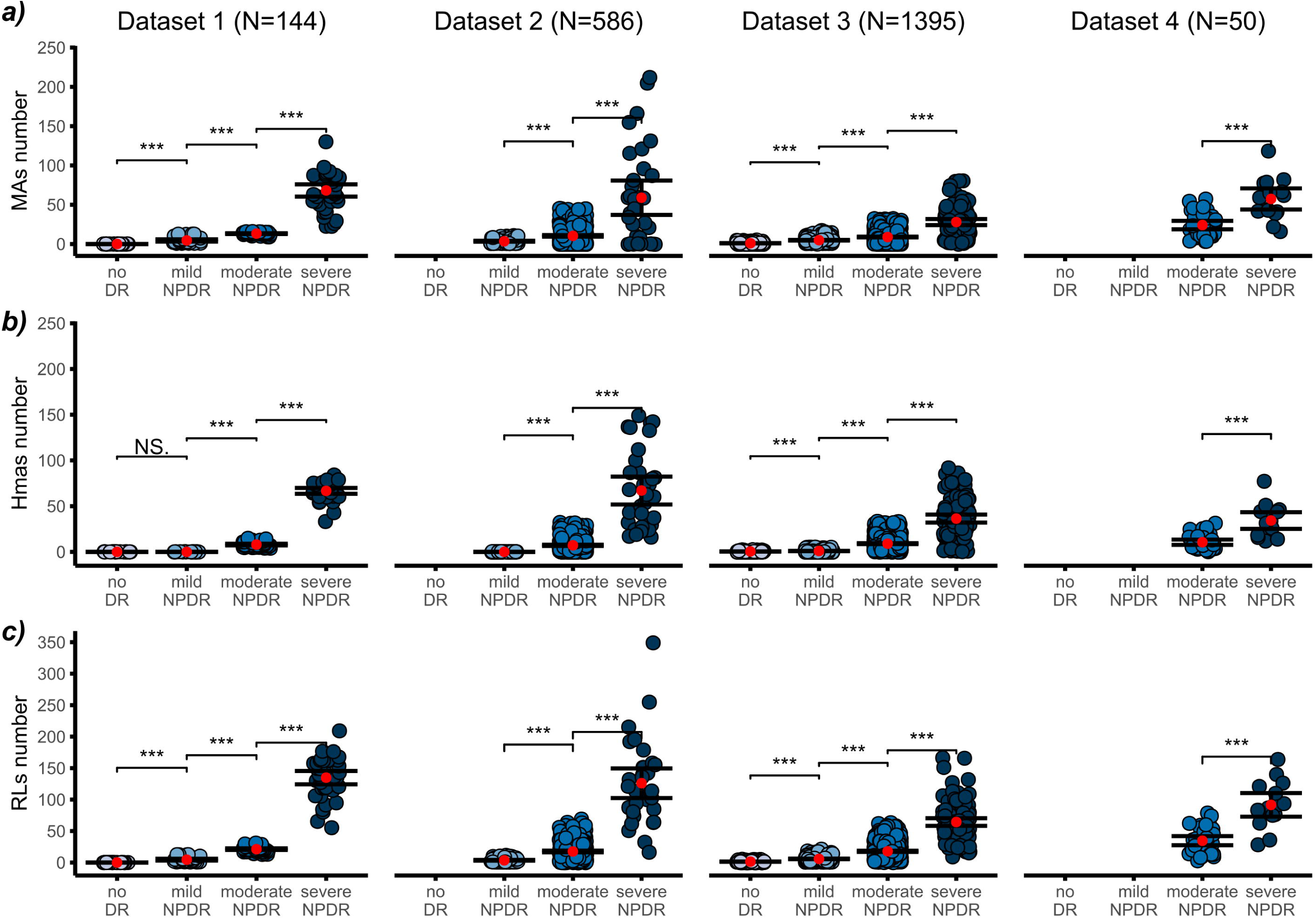
Number of lesions counted in the central retinal fundus image plotted against ICDR severity level (mean ±95% CI), for each dataset. a) number of microaneurysms (MAs); b) number of hemorrhages (Hmas) and c) number of red lesions (RLs). Level of significance: *** p<0.001; N.S. means not significant.

Remarkably, a substantial and statistically significant increase (p<0.001) in the prevalence of MAs was observed across all categories of the ICDR classification system within any of the datasets analyzed. Consistently, the mean MA values for no DR category remained negligible, aligning with the clinical definition of this particular stage, in those datasets incorporating MA segmentations at no DR level (Dataset 1 and Dataset 3). Globally, mild and moderate cases exhibited mean MA values of 4.3 (95% CI of 3.8 to 4.8) and 14.2 (95% CI of 12.8 to 15.5), respectively. In severe NPDR levels, MA counts escalated significantly, reaching an average of 53.1 (95% CI of 46.7 to 59.6), aligned with the expected dispersion in pre-proliferative patients. Despite the inherent variability associated with severe stages, attributed to its multifaceted clinical manifestations, statistical significance persisted, affirming the robustness of our observations.

A parallel trend is observed regarding the number of Hmas, with a statistically significant increase noted across NPDR severities (p<0.001), irrespective of the dataset analyzed. Noteworthily, within the Dataset 1, the change from no DR to mild NPDR lacks statistical significance, as the Hmas count remains identical (null) at both levels. However, in the context of Dataset 3, a marginal rise in Hmas numbers is noted in mild NPDR, yielding statistical significance. In both cases, our interpretation aligns with clinical guidelines and the ICDR definition,^27^ suggesting that Hmas are typically absent in both no DR and mild cases. In terms of actual counts, Hmas showed a very similar trend compared to MAs, with the mean count for moderate cases being 8.7 (95% CI of 7.9 to 9.4) and for severe cases 51.1 (95% CI 46.9 to 55.6).

Based on these findings, it is important to note that red lesions (RLs) also exhibited a comparable pattern to that observed with separate lesions, presenting a consistent trend across all datasets and high statistical significance (p<0.001). This observation holds particular value, considering the inherent challenge of distinguishing MAs and Hmas in fundus images. The mean values for RLs were negligible for no DR stage and 4.6 (95% CI of 4.1 to 5.2) for mild, 22.8 (95% CI of 21to 26.6) for moderate and 104.2 (95% CI of 96.5 to 111.9) for severe cases.

On the other hand, hard exudates (HEs) were analyzed separately due to their distinct vascular characteristics and propensity to cluster (Figure 2).

**Fig. 2.**
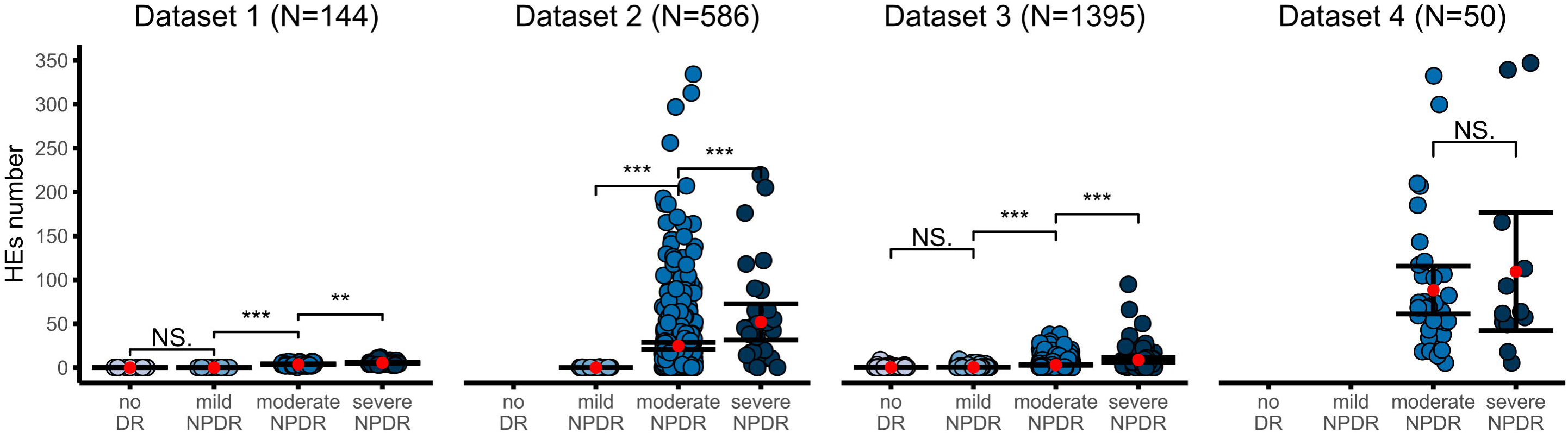
Number of Hard Exudates (HEs) in a single retinal field plotted against ICDR severity levels for each dataset (mean ±95% CI). Level of significance: *** p<0.001.

Interestingly, even though the quantification of HEs posed challenges owing to their tendency to aggregate into clusters,^28^ the resulting counts exhibited a statistically significant growth (p<0.001) along the DR severity scale, except for Dataset 4. Mean values were negligible for no DR and mild cases, but rose to 30 (95% CI of 23.3 to 36.6) for moderate and 58.2 (95% CI of 38.2 to 79.3) for severe NPDR. Again, the greatest dispersion of data was reported for Datasets 2 and 4, while the rest of databases maintained more tightly packed counts. This dispersion was further evidenced by the wider error bars observed for HEs compared to other lesions.

The overall analysis across datasets for MAs, Hmas, RLs and HEs in the rough population including prefiltered data led to the same conclusions, as the same pattern and the high statistical significance was maintained (p<0.001).

The data distributions illustrated in Figures 1 and 2 were also analyzed using weighted scores represented in a 3D plot, to simplify the examination of the principal parameter, the mean number of each retinal lesion, against the DR severity levels.

Figure 3 provides a comprehensive overview of lesion quantification across each DR severity level, in a 3D plot. The increases in lesion counts are consistent from no DR to mild and moderate categories and become more pronounced in severe NPDR cases, with higher impact on the RLs in the severe category. The tight confidence intervals evidence the reproducibility of lesion counts, proving that specific numerical values of MAs, Hmas and RLs can be reliably associated with DR severity levels. As previously noted, the dispersion of HEs is greater, particularly in severe cases, underscoring their variability. This global visualization also corroborates the clinical definitions of ICDR, with no DR category showing negligible number of lesions and mild NPDR lacking Hmas and HEs by definition.

**Fig. 3.**
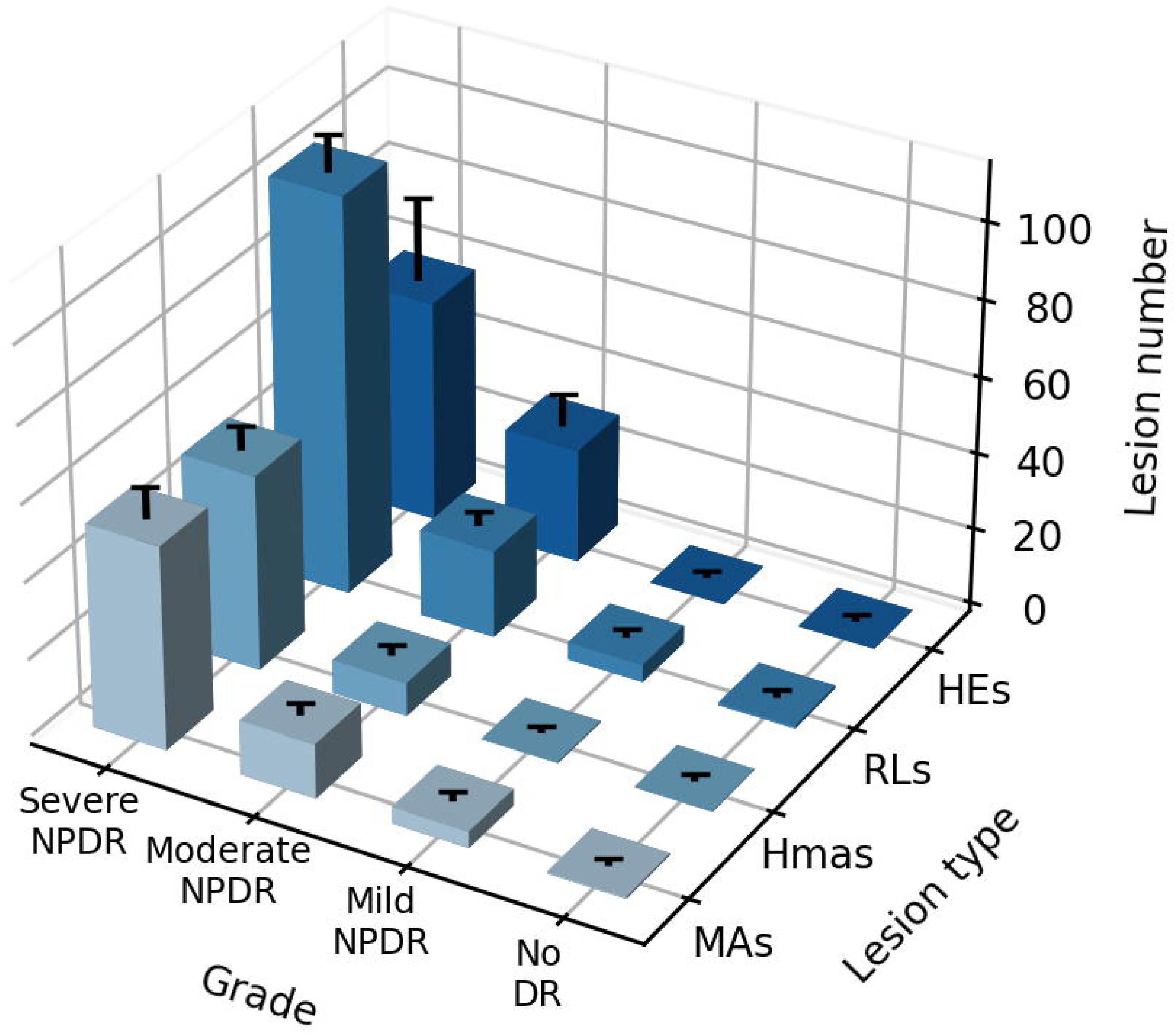
All-in-one 3D plot of weighted means (±95% CI) for MAs, Hmas, RLs and HEs (HEs) against ICDR severity levels.

Finally, we employed a classification neural network model trained on the standardized count values, extracted from the merged datasets to assess the predictive power of lesion counts. The data exhibits some class imbalance, with most cases falling within the moderate NPDR grades (Figure 4).

**Fig. 4.**
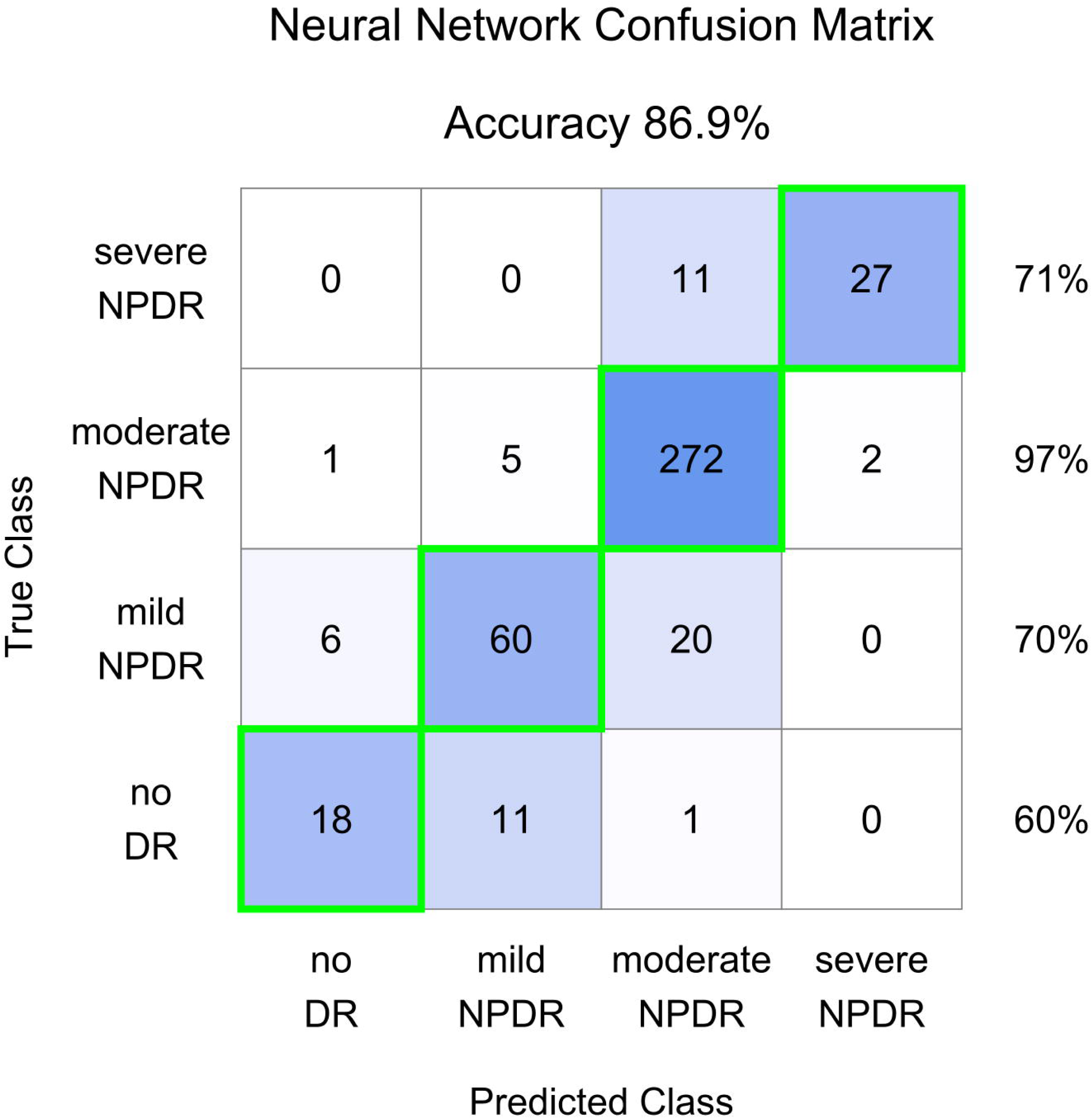
Confusion matrix for the neural network trained model outcome predictions. The right column represents the sensitivity (true positive rate) for each class: no DR, mild NPDR, moderate NPDR and severe NPDR.

Figure 4 illustrates the performance of the trained model, achieving an accuracy of 86.9%. Accuracy is notably high considering that predictions rely solely on one extracted feature (retinal lesion counts). By examining this feature from a single retinal image, it is possible to correctly diagnose most patients. Improved performance could be obtained through more data on the least represented groups.

The highest accuracy was achieved in moderate NPDR cases, with sensitivity over 95%. This category not only comprises the largest training sample, but also exhibits the most distinguishable traits associated with the retinal lesions under study. Moreover, a good true positive rate was obtained in both mild and severe NPDR, around 70%. Likely due to the scarce amount of trainable data on the ‘no DR’ class, the resulting predictions for this category slightly decreased to 60%.

## 4. Discussion

In this study, microaneurysms (MAs), hemorrhages (Hmas) and hard exudates (HEs) were automatically counted in a large set of lesion-segmented central retinal fundus images from different origins within a broad spectrum of Real-World Data (n=2,340), by using a watershed-aided algorithm, to corroborate the conclusions drawn from our previous study (n=60)^19^. The images used in this meta-analysis are mostly publicly available through open repositories of original and segmented (annotated) images.^17,19,24–26^ All non-proliferative stages have been represented, however, PDR has been excluded due to potential variations in the appearance and number of certain microvascular lesions in patients undergoing treatment. In fact, Esmaeilkhanian et al.^17^ reported a decrease on the number of lesions compared to the previous stage (severe NPDR).

The reporting of high statistical significance on the number of quantified lesions along DR severity levels across all databases analyzed (p<0.001), covering the four lesions under study, is particularly relevant, given the heterogeneity of images. In this regard, the fundus photographs of the diabetic population used vary in quality, field of view, focus and camera specifications, while efforts have been made to maintain the demographic diversity as extensively as possible. Despite these varied conditions, lesion quantification in a small central area of the retina has been demonstrated to significantly relate with the ICDR severity scale. This holds particular interest in the case of MAs, since this lesion has been widely reported as the first observable hallmark of DR, and their presence and extent in fundus images correlated with the risk of developing severe manifestations.^12,29^ If we further consider that their counting in a single central retinal field leads to a highly accurate classification for patient severity, we can confidently assert that MAs are a valid retinal biomarker for patient stratification in a quantitative fashion. This suggests that DR improvement or regression can be more closely evaluated and based on the frequency of relevant retinal lesions, as numerical metrics. Although Hmas followed the same trend and high statistical significance on their number among DR severity levels, they were more prone to confluence in moderate and severe stages, compromising the precision of segmentation. This observation leads to the conclusion that the nature of MAs, defined by punctate red dots, are more suitable for segmentation and quantification compared to other lesions, despite their significant statistical relevance.

Furthermore, it should be noted that analyses performed before data filtration led to the same levels of statistical significance (p<0.001). This observation reinforces the conclusions drawn from this study, demonstrating that despite using raw data collected under varied conditions and criteria, the outcomes remain unaltered. This finding is particularly compelling when considering that eyecare systems may follow different methods for capturing and processing images, yet the correlation remains robust.

RLs have also been explored. Besides showing high statistical significance (p<0.001), their detection in fundus images is easier compared to MAs and Hmas alone, making them a feasible option for current health systems where retina specialists still oversee image reading and grading. However, thanks to the progress of AI-assisted technologies, identification of individual lesions is poised to become a reality in the near future.^30–37^

Although HEs follow a statistically significant trend as well, their precise counting is challenging and prone to errors due to the intrinsic tendency of this lesion to aggregate, especially in severe forms.^28^ Consequently, the relationship between DR severity and the area occupied by HEs, rather than their count, has been studied (supporting Figure 1). It should be acknowledged that the utility of area measurement over count is only interesting in the most severe cases, which are characterized by large lesions. Otherwise, lesion number remains a solid metric for this purpose.

Despite of the strength of our findings, we acknowledge certain limitations in our study. Firstly, except for Dataset 1, all datasets were sourced from the public domain, where variations in DR grading criteria among different specialists may exist despite assuring the quality of grading and ground truths. In this line, the heterogeneity in image quality raised concern, due to the presence of out-of-focus photographs, cropped images or images with poor lighting, even though their contribution was not detrimental for the overall integrity and significance of the study results. Additionally, the scope of lesion types is constrained by the information publicly available, meaning that certain datasets only offer segmented lesions for specific severity grades (i.e. Dataset 4 includes segmented lesions only for moderate and severe NPDR graded stages), creating imbalanced categories. Finally, data refinement measures were implemented by discarding images with heavy atypical counts that were inconsistent with the harmonized clinical guidelines for defining the ICDR severity stages.

Secondly, a detailed examination was required on a subset of data from the Datasets 2 and 3 (n=37 out of 1981 images), as they included images graded as severe NPDR despite having a low number of MAs (<10), which contradicted our primary findings. By leveraging a supplementary neural network-driven tool, further insights into this new behavior were sought. Essentially, the analysis confirmed that images with a low MA count exhibited extensive hemorrhage covering all four quadrants, thereby justifying the Severe NPDR classification. However, this explanation did not apply uniformly across the entire subset, as some images also showed low counts for both MAs and Hmas. In such cases, a comprehensive analysis revealed the presence of intraretinal microvascular abnormalities (IRMA), which contributed to exacerbating the severity of DR.^27^

Expanding upon our insights, the relationship demonstrated between the quantification of several microvascular lesions and the ICDR severity underscores the feasibility of automated tools in refining diagnostic precision in DR. Such technologies offer the promise of enhancing the sensitivity and specificity of DR screening protocols, particularly in environments with limited access to specialized retinal care. Additionally, our results support the integration of quantitative analysis from a single fundus image in eyecare, which facilitates and improves diagnostic accuracy while providing clinicians with actionable insights at the point of care. The system described in this article additionally enables more thorough monitoring of patients diagnosed as mild NPDR, since they are often misdiagnosed or do not attend annual appointments due to their normal visual function. This approach allows for a higher attention to this patient group, as tracking the number of MAs, even low, and their changes over time can offer a clearer indication of risk of disease progression. While moderate to severe cases often receive more comprehensive monitoring due to the severity of DR, milder cases could be sometimes overlooked, so this method helps bridge this gap.

Systematic analysis of retinal images to quantify lesion counts enables more efficient patient management, and their risk of disease progression based on the number of MAs, can be individually assessed. These results remain consistent regardless of the camera specification, focus or procedural variations, ensuring that patients can be rapidly diagnosed and categorized in NPDR severity level, based on a single type of microvascular lesion. This approach not only aligns with the principles of precision medicine but also enhances the overall quality of care for patients with diabetes.

In conclusion, the quantification of microvascular lesions into a single central retinal field leads to a transformative step forward in the management of DR. Embracing this methodology could substantially reduce the burden of DR by facilitating more accurate diagnoses, timely interventions, and ultimately, improving the prognostic outcomes for millions of patients worldwide. This study reinforces the power of lesion counting to accurately determine the disease severity, and its integration into clinical protocols and consideration in clinical trials to efficiently monitor patient evolution relying on quantifiable individual microvascular lesions.

## Data Availability

All data produced in the present study are available upon reasonable request to the authors.

https://ieee-dataport.org/open-access/indian-diabetic-retinopathy-image-dataset-idrid

https://github.com/WeiQijie/retinal-lesions

https://www.kaggle.com/datasets/mariaherrerot/ddrdataset?resource=download

## Acknowledgements

The authors wish to thank the publicly available repositories of retinal images, as well as the retinal specialists who graded and annotated them. Data from Dataset 1 was kindly provided by the Instituto de Oftalmobiología Aplicada and published in a previous collaborative article.^19^

No funding was received for this research.

## Notes

### Competing Interest Statement

The authors have declared no competing interest.

### Funding Statement

This study was funded by BCN Peptides S.A.

### Author Declarations

The Research Ethics Committee (CEIm) of Valladolid East Health Area - HCUV issued a FAVORABLE STATEMENT on the research project regarding the proprietary data during the meeting held on 16/03/2023 (minutes no. 5 of 2023) and agrees that the said research project may be carried out by the principal investigator and their team. The publication of the image sets was authorized by the corresponding organizations before making them available. Informed consent was not required for the publicly available data.

