## supporting Figure 1 for "Microvascular Metrics on Diabetic Retinopathy: Insights from a Meta-Analysis of Diabetic Eye Images from Real-World Data"

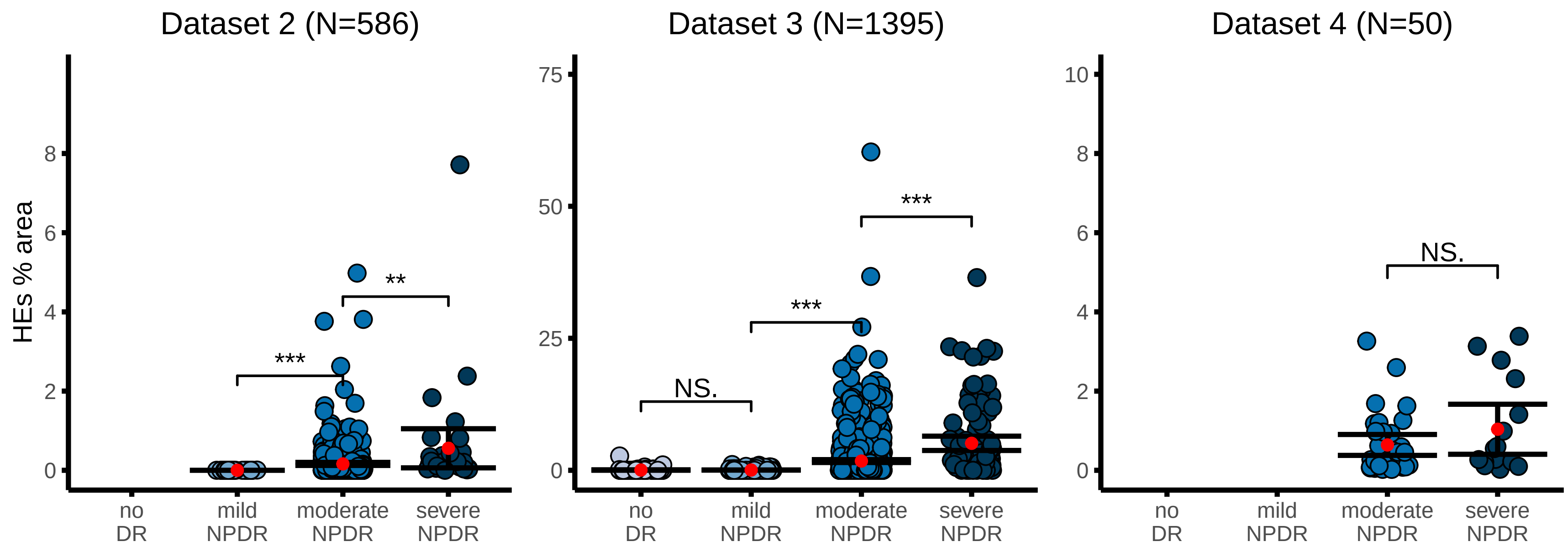


Figure 1: Percentage area of Hard Exudates (HEs) from a single retinal field plotted against ICDR severity levels for datasets 2,3 and 4 (mean ±95% CI). Level of significance: *** p<0.001; ** p<0.01; N.S. means not significant.
