## Supplementary material for "Microvascular Metrics on Diabetic Retinopathy: Insights from a Meta-Analysis of Diabetic Eye Images from Real-World Data": Table 1

### Data Summary for Lesion Types by Dataset

| | *Dataset 1* | *Dataset 2* | *Dataset 3* | *Dataset 4* |
|---|---|---|---|---|
| ***MAs number*** | | | | |
| *no DR* | 0 (±0) | | 1 (±0.2) | |
| *mild NPDR* | 4.6 (±1.5) | 3.5 (±0.6) | 4.8 (±0.4) | |
| *moderate NPDR* | 13.3 (±0.5) | 10.4 (±1) | 8.9 (±0.5) | 24.1 (±5.4) |
| *severe NPDR* | 68.1 (±7.8) | 58.9 (±21.9) | 27.9 (±3.9) | 57.4 (±13.4) |
| ***Hmas number*** | | | | |
| *no DR* | 0 (±0) | | 0.4 (±0.1) | |
| *mild NPDR* | 0 (±0) | 0 (±0) | 1 (±0.1) | |
| *moderate NPDR* | 8 (±0.9) | 7.2 (±0.7) | 9 (±0.5) | 10.6 (±2.9) |
| *severe NPDR* | 66.7 (3.2) | 67.1 (±15.3) | 36.4 (±4.4) | 34.2 (±9.1) |
| ***RLs number*** | | | | |
| *no DR* | 0 (±0) | | 1.4 (±0.2) | |
| *mild NPDR* | 4.6 (±1.5) | 3.5 (±0.6) | 5.8 (±0.4) | |
| *moderate NPDR* | 21.3 (±1.3) | 17.5 (±1.3) | 17.8 (±0.9) | 34.7 (±7.2) |
| *severe NPDR* | 134.8 (±10.6) | 126 (±23.5) | 64.3 (±6.1) | 91.6 (±18.8) |
| ***HEs number*** | | | | |
| *no DR* | 0 (±0) | | 0.3 (±0.2) | |
| *mild NPDR* | 0 (±0) | 0 (±0) | 0.4 (±0.1) | |
| *moderate NPDR* | 3.8 (±0.5) | 24.7 (±4) | 3 (±0.3) | 88.3 (±27.2) |
| *severe NPDR* | 5.4 (±0.8) | 52.1 (±20.7) | 8.7 (±2.6) | 166.7 (±84.5) |

**Table 1**. Mean (±95% CI) for each lesion type, severity grade and dataset, corresponding to data from Figures 1 and 2.
