## Supplementary material for "Microvascular Metrics on Diabetic Retinopathy: Insights from a Meta-Analysis of Diabetic Eye Images from Real-World Data": Table 2

### Weighted Data Summary for Severity Grades

| | ***MAs*** | ***Hmas*** | ***RLs*** | ***HEs*** |
|---|---|---|---|---|
| *no DR* | 0.5 (±0.1) | 0.2 (±0.1) | 0.7 (±0.1) | 0.2 (±0.8) |
| *mild NPDR* | 4.3 (±0.5) | 0.3 (±0.1) | 4.6 (±0.5) | 0.1 (±0.1) |
| *moderate NPDR* | 14.2 (±1.3) | 8.7 (±0.8) | 22.8 (±1.8) | 30 (±6.6) |
| *severe NPDR* | 53.1 (±6.5) | 51.1 (±4.5) | 104.2 (±7.7) | 58.2 (±6.3) |

**Table 2**. Weighted means (±95% CI) for each lesion type and severity grade, corresponding to data from Figure 3.
